# Religiosity and mental wellbeing among members of majority and minority religions: findings from Understanding Society, The UK Household Longitudinal Study

**DOI:** 10.1101/2020.02.25.20027904

**Authors:** Ozan Aksoy, David Bann, Meg E Fluharty, Alita Nandi

**Author notes:** Contributed equally (Ozan Aksoy, David Bann).

## Abstract

**Objectives:** To examine the associations between religious affiliation, religious service attendance, subjective religious beliefs and mental wellbeing among the religiously unaffiliated, Christians, Muslims, and members of other minority religions in the UK using a longitudinal design.

**Methods:** We used data from four waves (2009–2013) of the UK Understanding Society, a longitudinal household panel survey with over 70,000 individuals in 30,000 households which included 4,000 households from an Ethnic Minority Boost sample. We adjusted for potential confounders (including ethnicity, socioeconomic factors and personality) and accounted for household fixed effects that absorb unobserved confounding factors operating at the household level. Outcomes were the Shortened Warwick-Edinburgh Mental Wellbeing Scale (SWEMWBS) and the General Health Questionnaire (GHQ).

**Results:** Compared with Christians and the nonreligious, Muslims and members of other minority religions reported significantly lower mental wellbeing, indicated by higher SWEMWBS and lower GHQ scores. These differences were only partially accounted for by confounding factors, by ethnicity and by the mediators we examined. Amongst those with religious affiliations (Christians, Muslims, and others), higher religious service attendance was associated with higher SWEMWBS; amongst those with no religious affiliation, there was no association. Higher religious service attendance is associated with lower GHQ scores amongst those with and without religious affiliations. The subjective importance of religion was not associated with SWEMWBS yet was associated with higher GHQ scores.

**Conclusions:** Religious service attendance as opposed to the subjective importance of religion appears to have positive effects on mental wellbeing outcomes. This suggests that the positive effects of religion on mental health operate through social channels. Findings point to the potential benefit of secular alternatives to religious service attendance to improve population-wide mental wellbeing.

## Introduction

Mental health and wellbeing are important to individuals, families, and society. Mental ill health is a leading contributor to the global burden of disease^1^ which motivates a need to better understand its modifiable determinants. There is increasing awareness that mental health and wellbeing are multidimensional constructs; positive mental wellbeing may be a different construct to mental ill health.^2^ Across the non-disordered population, higher positive mental wellbeing appears to have protective effects on other important outcomes such as physical health,^3^ and socioeconomic outcomes such as productivity.^4^ It is therefore important to identify the modifiable determinants of both mental ill health and positive mental wellbeing.

A growing body of literature—largely conducted in the US^5^ and mostly cross-sectional in nature^6 7^—has suggested that greater religiosity (particularly religious service attendance) is associated with reduced mental ill health risk and greater subjective wellbeing. Religious attendance may benefit these outcomes through a myriad of mechanisms, such as by reducing loneliness, increasing social interaction and support, and fostering engagement with other community services.^6 8^ Conversely, there could be adverse effects, such as through feelings of guilt associated with some religious beliefs, or ostracization from other secular societal activities that might benefit wellbeing. Interpretation of the existing literature is however currently hampered by difficulty in generalizability—any effect of religiosity on wellbeing outcomes is likely to differ by societal context and religious denomination. Yet most existing work has been conducted exclusively in the US, analyzing effects of Christian religious attendance in a context in which Christian faith is a prominent part of public and political life.^9^ More research is therefore needed outside of the US, including analysis of other religious groups.^5^ Since associations between religiosity and outcomes may be due to confounding and/or reverse causality,^10^ research using alternative empirical strategies is also required.

We extend the existing literature by examining associations between multiple religiosity measures and wellbeing outcomes in the UK—a secular country compared with the US.^9^ We used Understanding Society, a large nationally representative household panel study which contains information on religious affiliation, attendance, and the perceived importance of religion. Its ethnically diverse sample contains considerable heterogeneity in each of these religiosity measures. We hypothesized that greater religious service attendance would benefit wellbeing across Christian and Muslim groups as well as members of other minority religions,^11^ yet average wellbeing would be lower among Muslims and members of other minority religions, due to their increased exposure to discrimination,^12 13^ socioeconomic disadvantage,^14^ and higher levels of acculturation stress.^14 15^ Finally, we used the household nature of the study to examine within-household differences in wellbeing outcomes in order to account for unobserved confounding at the household level.^16^ We hypothesized that effects of religious service attendance would be partly but not fully explained by such household-level confounders such as family socioeconomic status and shared heritable^17^ determinants of wellbeing.

## Methods

### Data

Data from Understanding Society (The UK Household Longitudinal Study, UKHLS) were used. UKHLS is a nationally representative household panel study which started in 2009 with over 70,000 individuals in 30,000 households which included 4,000 households from an Ethnic Minority Boost sample.^18^ In the 2^nd^ Wave of the study, 8,000 households from the long running BHPS sample were added. The study follows these original sample members (OSM) and attempts to interview 10+ year olds every year. Individuals who join these households are also interviewed only while they are co-resident with these OSMs. When interviewed face-to-face by an interviewer, questions on wellbeing and other sensitive information are collected via questionnaires that the respondent completes by themselves to reduce social desirability bias. Detailed study information and sampling methodology can be found elsewhere.^18^ All participants consent for use of their anonymised survey information, and data for this study was accessed through the UK Data Service (https://www.ukdataservice.ac.uk/).

The sample for our analysis includes responders of Understanding Society Survey who took part in Wave 1 (2009/2011) and Wave 4 (2012-2014) and have responded to questions on religiosity or wellbeing. We also use data collected in Wave 3 (2011-2013) as in this wave information was collected on personality traits and the number of close friends. As a result, the final sample was composed of individuals who responded in either of Wave 1, 3 and 4. Finally we use the outcome variables in Wave 2 (2010-12) for robustness checks.

### Religion and wellbeing measures

Religious affiliation and religiosity were captured in three ways. First, participants identified whether they belong to any religion and if so, which religion they belong to. We collapsed responses into four group: non-religious, Christian of any denomination, Muslim, and other minority religions (Sikh, Jewish, Buddhist, Hindu). Religious attendance was measured by asking ‘*How often, if at all, do you attend religious services or meetings?* with categorical responses of ‘*weekly’ ‘monthly’ ‘yearly’ ‘never or practically never’*, or *‘only at weddings, funerals etc*.*’* We swapped the final two categories, standardized it to be between 0 and 1. Finally, the importance of religion was captured by asking ‘*how much of a difference would you say religious beliefs make to your life*?’ with categorical responses of ‘*a great’, ‘some’, ‘a little’*, or *‘no’ difference*. This was similarly standardized to values between 0 and 1.

Wellbeing was measured using the Shortened Warwick-Edinburgh Mental Wellbeing Scale (SWEMWBS)^19 20^ and the General Health Questionnaire (GHQ).^21 22^ SWEMWBS is constructed to capture positive mental wellbeing in a unidimensional construct. Participants were asked seven questions corresponding to their feeling and thoughts in the past 2 weeks such as “I’ve been feeling optimistic about the future” and “I’ve been feeling close to other people”, with responses on a Likert scale ranging from “none of the time” to “all the time” (scores range from 7-35 with higher scores indicating better mental wellbeing). The GHQ is an affective or experienced measure of wellbeing/mental health capturing anxiety, stress and depressive symptoms. It includes 12 questions which ask how a person felt recently and each question is measure on a four-point Likert scale—items capture information on concentrate problems, sleep concerns, and difficulty in decision making (scores range from 0- 36 with higher scores indicating worse mental health).

### Potential confounders

The following were considered as potential confounders: age in years, gender (male/female; there was no evidence for religiosity × gender interaction), ethnicity (18 category measure), country of birth (England, Scotland, Wales, Northern Ireland, and non-UK), marital status, region (12 category Nuts-1 regions of the UK), education (degree, other degree, A-levels, GCSE, other qualification, no qualification), employment status (in paid employment or not), income (total net personal income), personality traits (Big 5), self-rated general health, natural logarithm of the number of close friends + 1, and the extent to which the respondent talks regularly to neighbours.

### Analytical Strategy

Associations between religiosity (wave 1) and wellbeing outcomes at wave 4 were examined using linear regression models. We selected outcomes from wave 4 to help reduce bias due to reverse causality. We also examined associations with outcomes at wave 2. Since potential some confounders (e.g., income, self-rated health) may operate as mediators in the links between religiosity and mental wellbeing outcomes, sequential adjustments were made to aid interpretation. Models were first fitted only with the three religion variables, then adjusted for 1) potential confounders (age, sex, ethnicity, country of birth); 2) additionally adjusted for the outcome variable measured in wave 1; and 3) additionally adjusted for potential confounders and/or mediators (self-rated health, marital status, income, region, personality, education, friends, and communication with neighbors). All linear regression models were conducted using Full Information Maximum Likelihood estimation (FIML) to reduce the impact of missing data on power and potential bias (findings were similar using complete case analyses).

We then used multi-level models to control for households fixed effects;^16^ these models account for unobserved confounders which operate at the household level, such as unmeasured socioeconomic or cultural factors. To help illustrate interpretation, such models estimate the mean differences in wellbeing outcomes of more religious persons in each household compared with the average household level. Robust standard errors were used in these models.

### Additional and sensitivity analyses

To examine whether mean differences in outcomes by religiosity groups was due to differences in the lower or upper tails of the wellbeing distributions, we examined associations between religiosity and wellbeing using quantile regression models. To examine whether wellbeing outcomes were comparable across religiosity groups, we checked for measurement invariance across religious affiliation groups and identified potentially problematic items. Main analyses were then repeated removing these potentially problematic items. To examine if findings were similar across an alternative wellbeing outcome, we repeated main analyses using self-reported life satisfaction (ranging from 0-7) as an outcome. Since ethnicity may be an important confounder—yet may be co-linear with religious affiliation and thus lead to over-adjustment bias—we repeated analyses without adjusting for ethnicity. Finally, we conducted analyses before and after applying survey design weights to examine if this altered main findings.

## Results

25,114 participants had complete data for religious affiliation, importance of religion, service attendance, and mental health/wellbeing outcomes in wave 4. 61,462 participants have non-missing data for at least one outcome variable or covariates. See Table 1 for sample sizes for each variable. GHQ and WEMWBS scores were strongly negatively correlated (-.65 in wave 4, -.61 in wave 1). As anticipated, those with Christian or Muslim religious affiliations were more likely than non-religious participants to report religion as being important and regularly attend religious practices (Figure 1). Muslims were more likely than Christian to report religion as being important and to regularly attend religious practices (Figure 1).

**Table 1.** Descriptive statistics of the study sample—Understanding Society Waves 1 (2009/2011) and 4 (2012- 2014).

|  | n | mean | sd | min | max |
| --- | --- | --- | --- | --- | --- |
| Wellbeing outcomes |  |  |  |  |  |
| SWEMWBS (wave 4) | 39135 | 24.7 | 4.54 | 7 | 35 |
| SWEMWBS (wave 1) | 38395 | 25.2 | 4.54 | 7 | 35 |
| GHQ (wave 4) | 38852 | 11.0 | 5.60 | 0 | 36 |
| GHQ (wave 1) | 39700 | 11.0 | 5.36 | 0 | 36 |
| Life satisfaction (wave 4) | 38908 | 5.06 | 1.53 | 1 | 7 |
| Life satisfaction (wave 1) | 39558 | 5.25 | 1.46 | 1 | 7 |
| Religiosity |  |  |  |  |  |
| Importance of religion | 47583 | 0.42 | 0.40 | 0 | 1 |
| Religious practice | 47674 | 0.39 | 0.34 | 0 | 1 |
| Religious affiliation |  |  |  |  |  |
| Nonreligious | 47659 | 0.44 | 0.50 | 0 | 1 |
| Christian | 47659 | 0.43 | 0.50 | 0 | 1 |
| Muslim | 47659 | 0.079 | 0.27 | 0 | 1 |
| Other religion | 47659 | 0.051 | 0.22 | 0 | 1 |
| Ethnicity |  |  |  |  |  |
| British/english/scott/welsh/NI | 47678 | 0.75 | 0.43 | 0 | 1 |
| Indian | 47678 | 0.040 | 0.20 | 0 | 1 |
| Pakistani | 47678 | 0.030 | 0.17 | 0 | 1 |
| Country of birth |  |  |  |  |  |
| England | 50978 | 0.66 | 0.47 | 0 | 1 |
| Scotland | 50978 | 0.070 | 0.26 | 0 | 1 |
| Wales | 50978 | 0.042 | 0.20 | 0 | 1 |
| N.Ireland | 50978 | 0.040 | 0.20 | 0 | 1 |
| Non-UK | 50978 | 0.19 | 0.39 | 0 | 1 |
| Education |  |  |  |  |  |
| Degree | 46937 | 0.23 | 0.42 | 0 | 1 |
| Other degree | 46937 | 0.11 | 0.32 | 0 | 1 |
| A-level | 46937 | 0.21 | 0.41 | 0 | 1 |
| GCSE | 46937 | 0.21 | 0.41 | 0 | 1 |
| Other | 46937 | 0.095 | 0.29 | 0 | 1 |
| No qualification | 46937 | 0.14 | 0.35 | 0 | 1 |
| General health | 47130 | 2.40 | 1.12 | 0 | 4 |
| Age | 50994 | 45.6 | 18.2 | 14 | 101 |
| Net income | 50994 | 1199.0 | 1226.2 | -12495.48 | 15000 |
| Personality (Big 5) |  |  |  |  |  |
| Agreeableness | 40625 | 5.63 | 1.04 | 1 | 7 |
| Conscientiousness | 40618 | 5.46 | 1.12 | 1 | 7 |
| Extraversion | 40626 | 4.59 | 1.30 | 1 | 7 |
| Neuroticism | 40626 | 3.57 | 1.44 | 1 | 7 |
| Openness | 40586 | 4.54 | 1.32 | 1 | 7 |
| Female | 75131 | 0.37 | 0.48 | 0 | 1 |
| Marital Status (selected categories) |  |  |  |  |  |
| Single | 47046 | 0.23 | 0.42 | 0 | 1 |
| Married | 47046 | 0.51 | 0.50 | 0 | 1 |
| Cohabiting | 47046 | 0.11 | 0.32 | 0 | 1 |
| Talks to neighbors (1 strongly agree to 5 strongly disagree) | 39337 | 2.29 | 1.00 | 1 | 5 |
| Log(# close friends + 1) | 45385 | 1.62 | 0.63 | 0 | 4.62 |
| Employed | 75131 | 0.31 | 0.46 | 0 | 1 |

**Figure 1.**
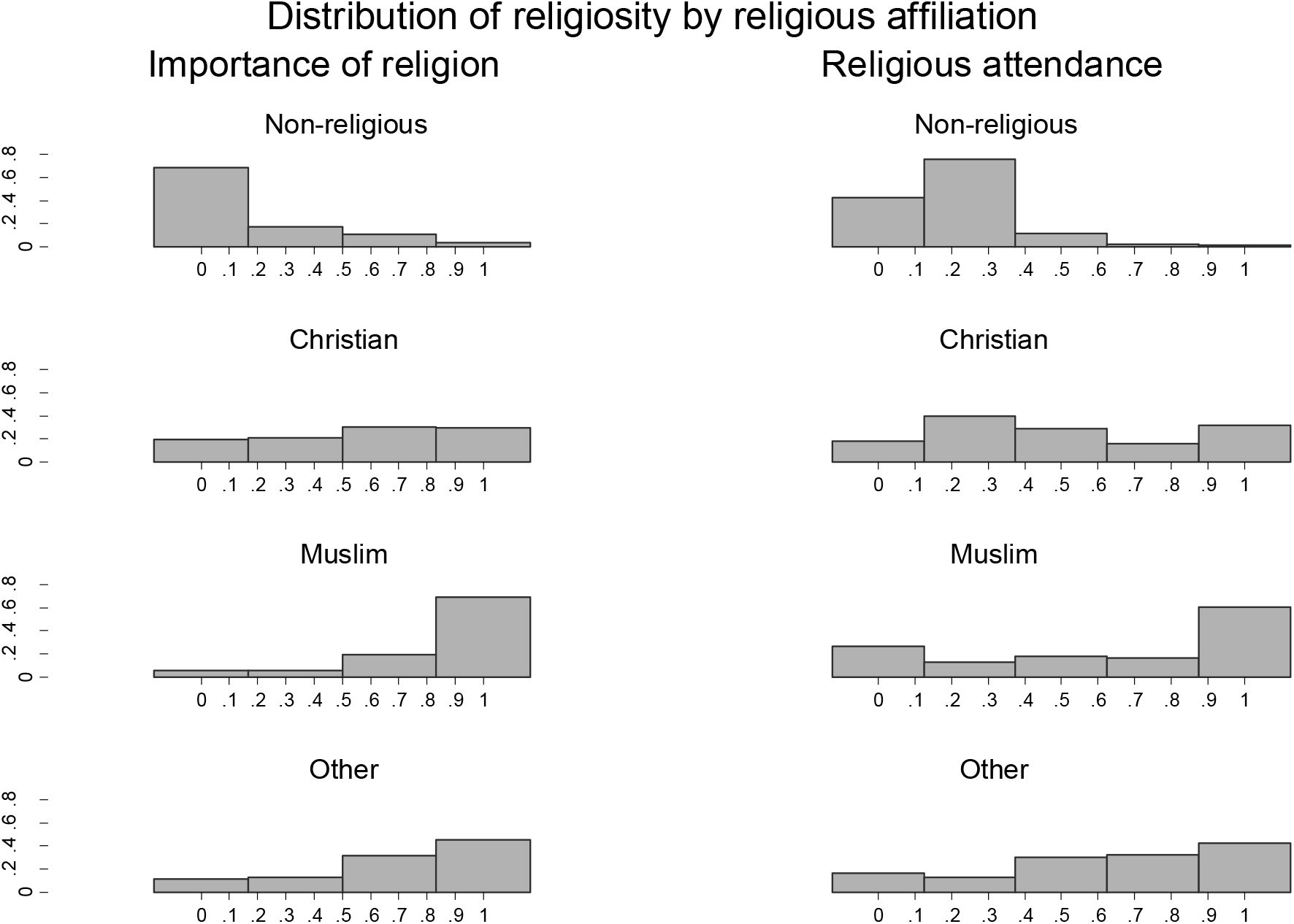
Distributions of perceived importance of religion and religious service attendance by religious affiliation. Note: percentages shown on Y-axes.

### Religious affiliation and mental health/wellbeing outcomes

Muslims and members of other minority religions had higher average GHQ and lower SWEMWBS scores than non-religious and Christian participants (Figures 2 and 3). Christians had higher SWEMWBS scores than the non-religious. These differences attenuated to null after adjustment for potential confounders and mediators, but the SWEMWBS difference between Muslims and the non-religious remained albeit attenuated after this adjustment (Figure 2 and Figure 3). This attenuation was particularly driven by ethnicity for Muslims and age for Christians. Because members of a household are generally of the same religion, models with household fixed effects do not result in meaningful estimates of the coefficients for religious affiliation.

**Figure 2.**
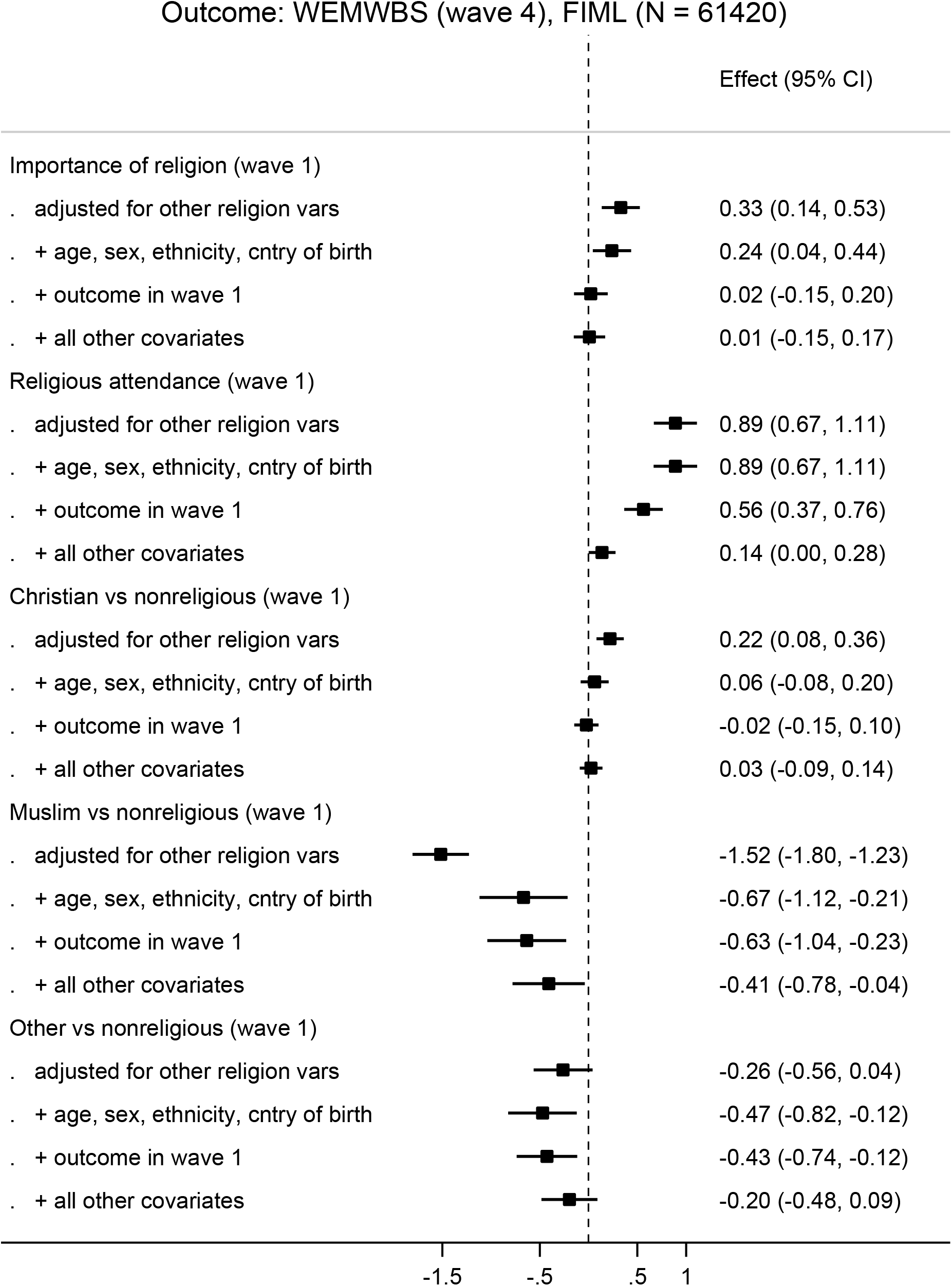
Associations between religiosity measures and positive mental wellbeing (WEMWBS scale). Note: Wave 1 (2009/2011) and Wave 4 (2012-2014); Full Information Maximum Likelihood Estimation (FIML) was used to account for missing exposure and confounder data.

**Figure 3.**
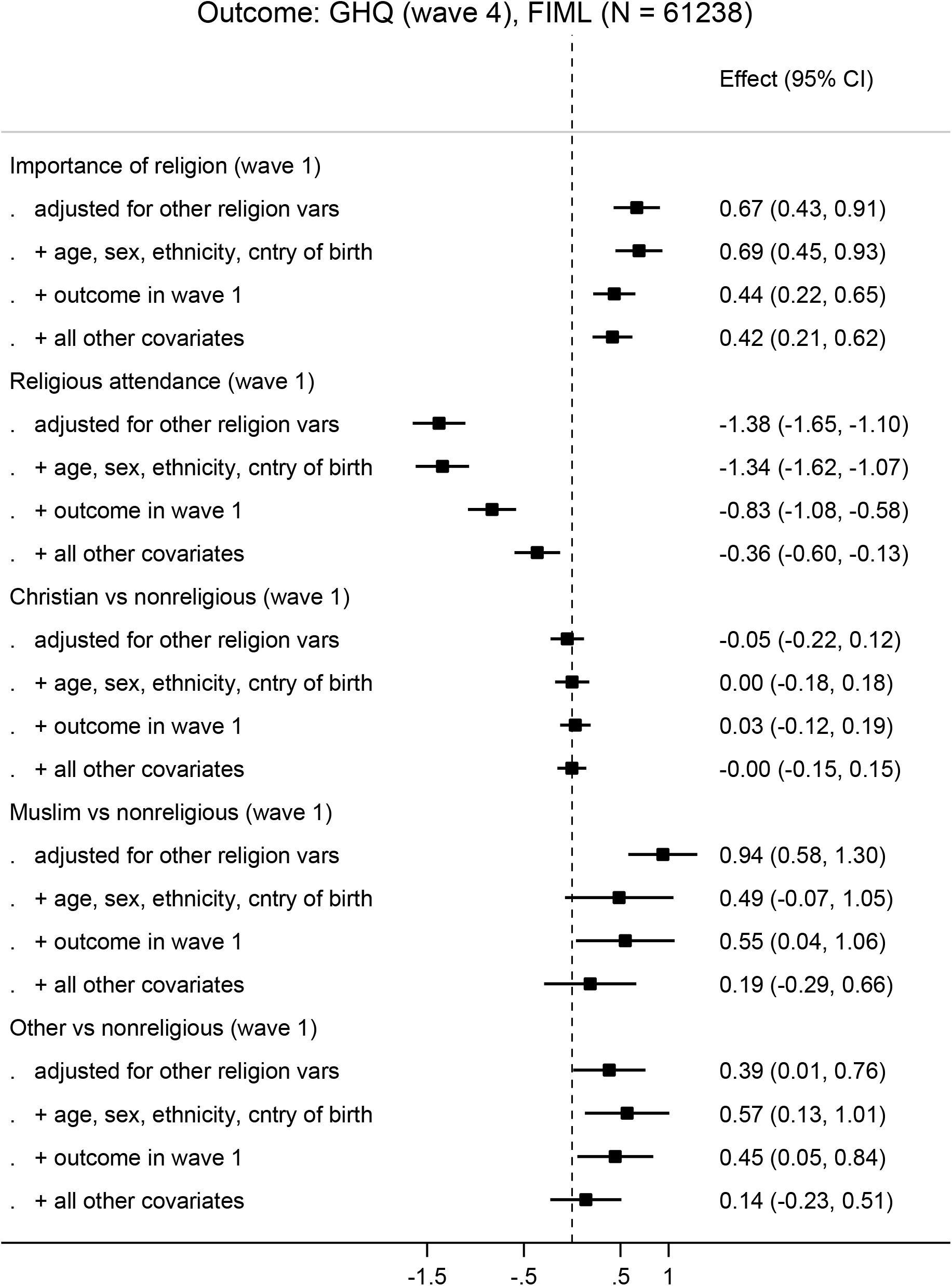
Associations between religiosity measures and negative mental wellbeing (General Health Questionnaire (GHQ) score). Note: Wave 1 (2009/2011) and Wave 4 (2012-2014); Full Information Maximum Likelihood Estimation (FIML) was used to account for missing exposure and confounder data.

### Importance of religion and mental health/wellbeing outcomes

Higher reported importance of religion was associated with higher SWEMWBS and GHQ scores (Figures 2-3). The association with SWEMWBS attenuated to null once a minimal set of confounders were accounted for (Figure 2); when household fixed effects were accounted for, the association switched sign, but 95% confidence intervals included the null (Figure 4). The association with GHQ remained even after controlling for potential confounders and mediators (Figure 3), and when accounting for household fixed effects (Figure 4).

**Figure 4.**
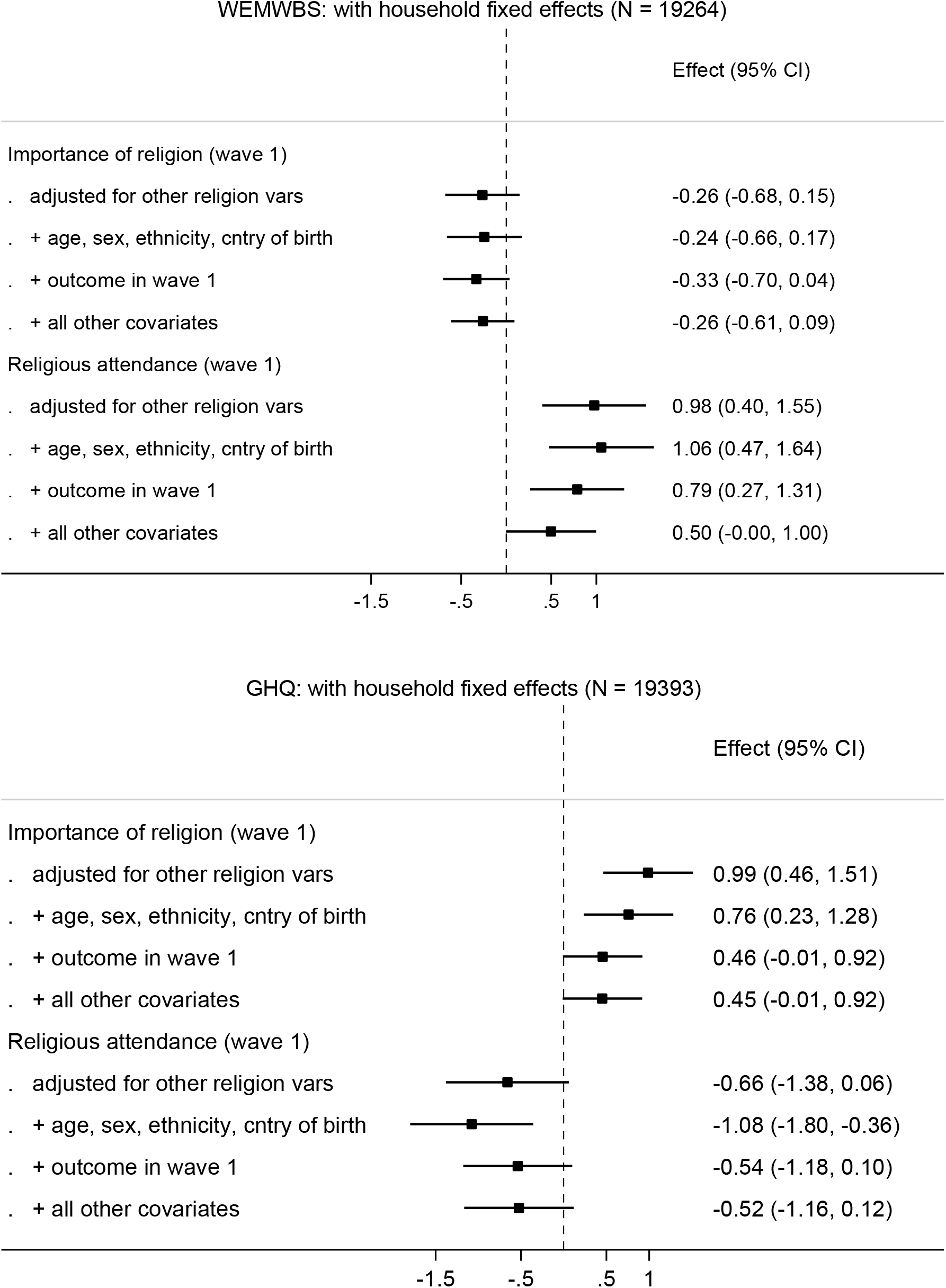
Associations between religiosity measures and mental wellbeing outcomes, accounting for household fixed effects. Note: Religiosity measured in Wave 1 (2009/2011) and outcomes in Wave 4 (2012-2014).

### Religious service attendance and mental health/wellbeing outcomes

Religious service attendance was associated with higher SWEMWBS and lower GHQ scores (Figures 2-3). For example, there was a 0.89 (95% CI: 0.67, 1.11) difference in SWEMWBS score comparing those with the most compared with least religious attendance (Figure 2). This change corresponds to about 20% of the standard deviation in SWEMWBS scores (Table 1). These differences were still found, albeit partly attenuated, after adjustment for potential confounders and mediators (Figures 2-3). After accounting for household fixed effects, associations were similar, with stronger evidence for associations with SWEMWBS than GHQ (Figure 4).

As anticipated, the association between attendance and SWEMWBS was stronger for those with Christian or Muslim affiliations comparted with no religions (Figure 5); there was no association amongst those with no affiliation. There is no evidence in the data that the association between attendance and GHQ varies by religious affiliation (*P* = 0.946).

**Figure 5.**
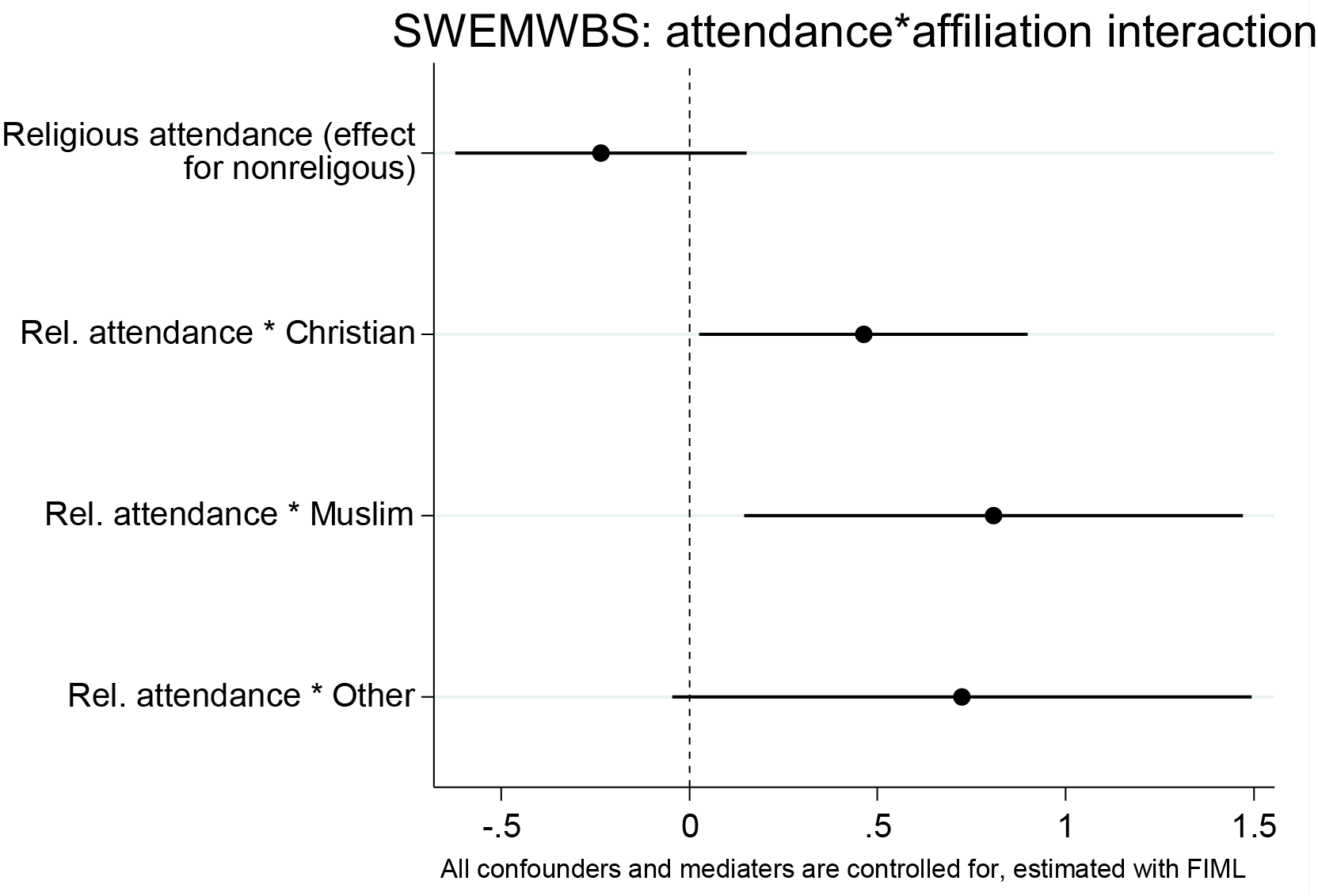
Associations between religious service attendance and mental wellbeing, including an interaction of religious service attendance x religious affiliation. Note: Religious attendance measured in Wave 1 (2009/2011) and outcomes in Wave 4 (2012-2014).

### Additional and sensitivity analyses

Quantile regression analysis suggests that the average differences reported above were driven particularly by differences at the lower (most negative) parts of the mental health/wellbeing distribution—that is, the lower end of the SWEMWBS distribution and the higher end of the GHQ distribution (Figure 6). This shows that while the average associations between religion variables and mental wellbeing reported above appear small (Figures 2-4), for the most negative parts of the mental wellbeing distribution the associations are rather substantial.

**Figure 6.**
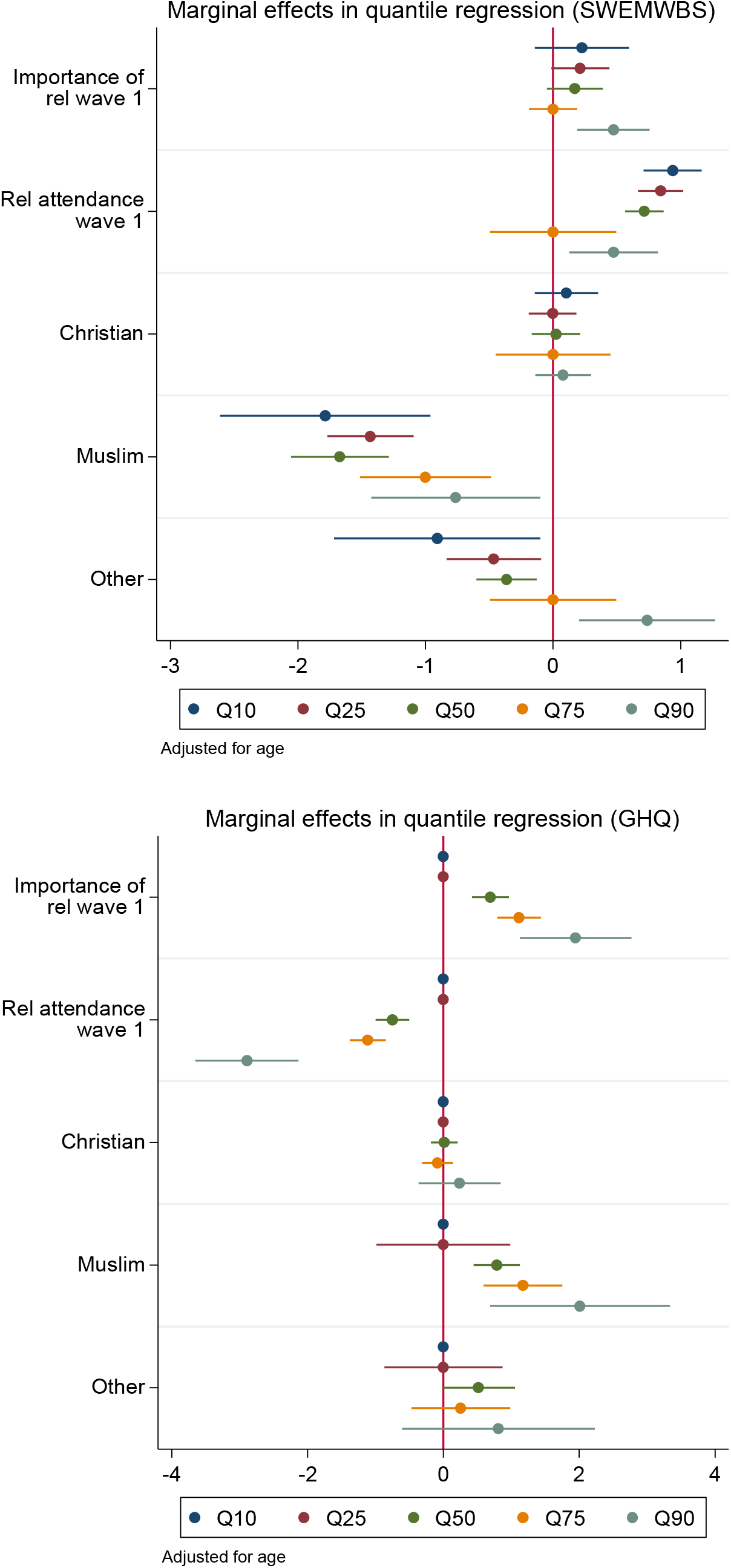
Associations between religiosity measures and mental wellbeing outcomes. Note: Religiosity measured in Wave 1 (2009/2011) and outcomes in Wave 4 (2012-2014); quantile regression models were used—coefficients are interpreted analogously to linear regression: e.g., Q50 shows the median difference in mental wellbeing comparing those with Muslim compared with no religious affiliation, while Q90 shows the difference at the 90^th^ quantile (far right-end of the distribution).

Findings were similar when 1) excluding items which lowered psychometric invariance of SWEMWBS and GHQ (Supplementary Figure A3); 2) using life satisfaction as an alternative outcome (Supplementary table D1 and D2); 3) not adjusting for ethnicity (Supplementary Table C1); and 4) applying survey weights (Supplementary Table B1); and 5) regressing health outcomes in wave 2 on religion variables in wave 1 (Supplementary Figure E1).

## Discussion

### Main findings

Using nationally representative household data from the UK, we found that Muslims and members of other minority religions had lower average mental wellbeing scores than Christians or those with no religious affiliation. These findings were present across two different outcomes and were only partially accounted for by potential confounding or mediating factors.

For those with religious affiliations (Christians, Muslims, and others), we found that higher religious service attendance was associated with higher mental wellbeing and lower common mental health symptoms. Interestingly, this positive effect was absent for those with no religious affiliation. These findings were robust to adjustment for multiple confounders and after accounting for household fixed effects. Quantile regression analysis suggested that the potential beneficial effects of service attendance were largely driven by those at particularly low levels of wellbeing. In contrast, the subjective importance of religion was not associated with higher mental wellbeing—in fact it was associated with higher mental health symptoms.

### Comparisons with previous evidence and explanation of findings

Our findings are consistent with previous evidence—largely conducted in the US on samples of Christians, suggestions of beneficial effects of religious attendance.^5 6 7^ Findings are also consistent with the only randomized controlled trial to which we are aware, suggesting causal effects of religiosity.^23^ However, given the specific intervention (evangelical Christian) and target population (low-income Filipino households), generalizing from this trial is challenging. Findings are also in line with a natural experimental study suggesting that greater involvement in a religious activity (Ramadan fasting) has a causal effect on higher wellbeing amongst Muslims, despite also being correlated with lower economic performance.^11^

We observed notable differences in wellbeing according to religious affiliation. Recent research has shown that globally Christians are happier than the nonreligious and Muslims, and the nonreligious and Muslims have similar levels of life satisfaction.^24 25^ In the UK, however, the differences between Christians and the nonreligious is practically nil once we account for a minimal set of covariates. Moreover, members of minority religions have significantly lower levels of wellbeing than Christians and the nonreligious. This suggest that the association between religious affiliation and wellbeing is context dependent, and members of minority religions are at risk of having lower mental wellbeing even in a highly secular country like the UK.

Taken together, our results are consistent with there being a positive causal effect of religious service attendance as opposed to subjective religious beliefs on mental wellbeing.^8^ The effect of service attendance may operate via multiple mechanisms, which may differ depending on the religion and societal context. These mechanisms include direct and indirect impacts of social networks such as social and community support, reducing loneliness, and fostering engagement with other community services.^8^ We found that (after adjusting for service attendance), greater reported importance of religion had no or potentially negative associations with wellbeing. This finding may reflect guilt associated with some religious beliefs or the stress of functioning in a secular society where some religious doctrines may not coincide with secular principles of the society.

The negative associations between belonging to a minority religion and mental health may reflect harassment and discrimination,^12 26^ socioeconomic disadvantage,^14^ and higher levels of acculturation stress^14 15^ that those with minority religions are more likely to experience. Religious service attendance, however, is positively associated with wellbeing among members of minority religions. This suggest that, consistent with past research, service attendance may buffer the negative consequences of belonging to a minority religion.^27^

Despite our use of longitudinal data, accounting for multiple potential confounders and household fixed effects, our findings may still reflect non-causal relationships. First, findings may reflect reverse causality—mental ill health may impede attendance in religious activities. While we used longitudinal data, adjusted for baseline mental ill health/wellbeing scores, there may be remaining residual impacts of preceding mental health on the religious attendance. Reverse causality may also impact on analysis within households (fixed effect analysis), yet this method is likely to better account for time invariant confounding factors such as family socioeconomic status. Ultimately, given the practical and ethical barriers to using randomized trials in this topic—and difficulties in generalization from trials which do exist—inference is guided by findings from observational studies such as the present one.

### Strengths and limitations

Our study was limited by a relatively short follow-up period (about 3 years). Thus, longer follow-up is required given concerns over reverse causality. Nevertheless, it is possible that causal beneficial effects of religious attendance are in fact short term in nature, and thus weak or non-existent when using longer periods of follow-up.

Strengths of the study include the large sample from a variable population, enabling examination of Muslims— a previously understudied group in studies of religion and wellbeing. Indeed, religion is noted stratifier of health inequality according to World Health Organization guidance.^28^ However, we should note that there is substantial within-group heterogeneity in each religion in terms of religious belief and practice which we were unable to investigate. While we used a large nationally representative study with considerable religious heterogeneity, we were underpowered to investigate wellbeing outcomes in smaller religious groups. This warrants investigation in studies with more granular data on religious affiliation.

Our analyses also contained substantial data on potential confounders (e.g. personality, contact with neighbors, number of friends) which were not available in much previous research. We also use household fixed effects to control for all factors that are invariant at the household level. Hence, while residual confounding cannot fully be ruled out, controlling for confounding is one of the strengths of the current study. We also considered several outcome variables, mental wellbeing, mental distress, and subjective wellbeing rather than relying on single outcome measure. Reassuringly, our findings were broadly similar across both outcomes.

### Potential implications

If associations between religious service attendance and outcomes are causal in nature, our findings may have implications for strategies to improve population-wide mental health. Given the increasing levels of mental ill health observed in the population^29^ and the decline in religious attendance observed in the West,^9^ one naive suggestion would be that religious service attendance should be increased across the entire population. However, we would caution against such suggestions, since alignment is clearly required between individuals’ faith and the religious services available. Further, there may be other deleterious consequences of such attendance which we do not observe.^30^ Indeed, we found that associations between religious service attendance and positive wellbeing outcomes were limited to those with religious affiliations. Instead, out findings point to a need for secular alternatives to religious services which can replicate and/or improve upon its potential benefits, regardless of religious faith. Indeed, one potential explanation of the worsening of mental health outcomes in recent decades^29^ is the increasing individualization nature of society, characterized by declines in communal activities such as religious service attendance.^31^

Amongst those with religious faith, our findings may suggest that facilitating religious service attendance may be one means by which the negative consequences of belonging to a minority religion could be averted. Such considerations may particularly benefit already vulnerable groups—for example, a recent report implied that in the UK women’s access to several masjids could be improved by providing more facilities for women.^32^

### Conclusions

Several studies have suggested that greater religiosity, particularly religious service attendance, is associated with reduced mental ill health risk and greater subjective wellbeing. These studies were largely cross-sectional and mainly conducted in the US with a focus on Christians. Our study suggests that such associations are also present in the UK—for both Christians and members of minority religions including Muslims. Moreover, such associations were found using longitudinal data in which multiple potential confounders were accounted for, as well as household fixed effects. In addition, we found a strong negative association between belonging to a minority religion and mental wellbeing. Taken together, our findings suggest that secular alternatives to religious service attendance may improve population-wide mental health.

## Data Availability

Data used in the study is publicly available from the UK data archive (https://www.ukdataservice.ac.uk/). Replication material (analysis codes) are available upon request.

## Supplementary material

### A: Testing measurement invariance of SWEMWBS and GHQ across religions

#### Measurement invariance of SWEMWBS across religions

Here, we test measurement invariance of the 7-item version of the Warwick-Edinburgh mental well-being scale across three religion categories: the non-religious, Christians of any denomination, and Muslims of any denomination. We will use the data from Understanding Society Wave 1. The seven items are (1) feeling optimistic about the future (2) feeling useful (3) feeling relaxed (4) dealing with problems well (5) thinking clearly (6) feeling close to others (7) able to make up own mind. A graphical representation of the measurement model is given below.

**Figure A1.**
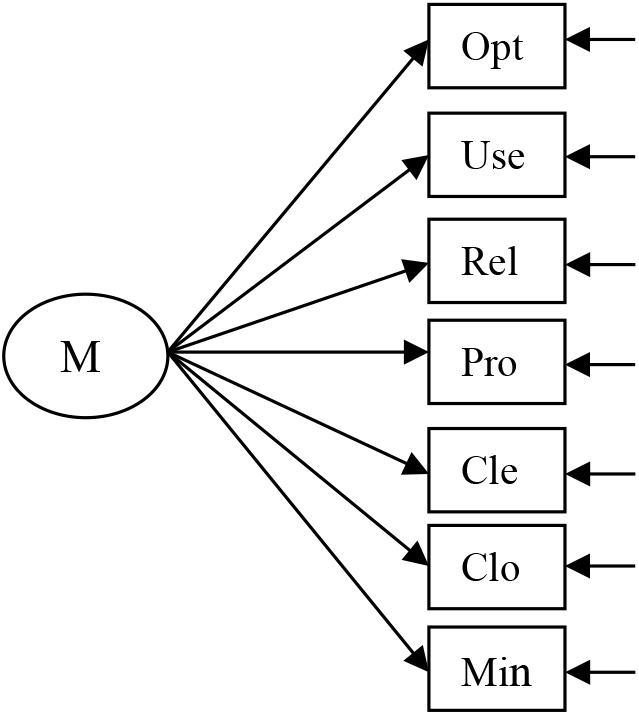

We fit four models with varying degrees of strictness of measurement invariance (see e.g. Kline 2016). The first one is the *configual invariance* model, which keeps the model structure in Figure X1 above the same, but allows the parameters to vary across the three groups. The second is the *weak invariance* model, which in addition to the configural invariance, constrains the loadings of the items on the latent mental wellbeing factor to be the same in the three groups. The third is a *strong invariance* model which constrains, in addition to the loadings, the intercepts of items to be the same across the three groups. Finally the *strict invariance* model constrains loadings, intercepts, and the error variances of the items.

Table A1 below shows a number of fit measures for the four models. Firstly, the exact fit hypothesis for the configural invariance is rejected. This implies that the most unconstrained model fits data less than perfect to start with. But also note that N is rather large in our case (36,623) so even small misfits may become insignificant. Our aim is not to test or validate the SWEMWBS scale itself, but establish its invariance across the three groups. So we leave a side the misfit of the configural invariance model. We would like to report, however, that adding a covariance between the error terms of item 1 and 2 and between item 5 and 7 improves the model fit rather significantly. Adding these two error covariances reduces the model χ2 to 2268.22 (36), RMSEA to 0.071, BIC to 556484.48, SRMR to 0.023 and increases CFI to 0.979. Because invariance of error covariances across the groups is not strictly required for sufficient measurement invariance, we carry on with the original specification without the error covariances. The conclusions reported below are the same if we carry out our analyses with the two error covariances added.

**Table A1:** Fit measures of various measurement invariance models; N = 36,623.

|  | Configural inv. | Weak inv. | Strong inv. | Strict Inv. |
| --- | --- | --- | --- | --- |
| $\chi^2$ | 6288.96*** | 6341.98*** | 6555.05*** | 6976.70*** |
| (df) | (42) | (54) | (66) | (80) |
| RMSEA | 0.110 | 0.098 | 0.090 | 0.084 |
| (90% CI) | (0.108-0.113) | (0.096-0.100) | (0.088-0.092) | (0.082-0.086) |
| AIC | 559906.14 | 559935.16 | 560124.23 | 560517.88 |
| BIC | 560442.17 | 560369.09 | 560456.06 | 560730.59 |
| CFI | 0.941 | 0.940 | 0.938 | 0.934 |
| TLI | 0.911 | 0.930 | 0.941 | 0.948 |
| SRMR | 0.039 | 0.046 | 0.052 | 0.068 |

The fit of the weak invariance model is comparable to the configural invariance model. While a likelihood ratio test favours the configural invariance over weak invariance (compare the χ2 values of the two models), this could again be due to very large N. Other fit measures, in fact, indicate that the weak invariance model fits somewhat better than the configural invariance model (e.g. RMSEA, BIC, and TLI). A comparison of item loadings across the three religion groups in the configural invariance model shows that the loadings vary only marginally. The largest differences are for the loadings of item 6 and 7, both of which are somewhat higher among Muslims and Christians than the non-religious. In fact, removing items 6 and 7 from the scale makes the χ2 difference test between the weak invariance and the configural invariance models statistically insignificant. While there is a case to remove these two items from the model, the sizes of the differences in the loadings across the religion groups seem rather minor.

The conclusions are similar if we compare the strong and strict invariance models with the configural invariance model. We thus conclude that the short version of the Warwick-Edinburgh mental well-being scale measures mental well-being rather similarly across the three religion groups with a caveat regarding item 6 and 7.

#### Measurement invariance of GHQ across religions

Now, we test measurement invariance of the 12-item subjective well-being score (GHQ) across the non-religious, Christians of any denomination, and Muslims of any denomination. We will use the data from Understanding Society Wave 1. The 12 items ask about (1) concentration (2) loss of sleep (3) playing a useful role (4) capable of making decisions (5) constantly under strain (6) problem overcoming difficulties (7) enjoy day-to-day activities (8) ability to face problems (9) unhappy or depressed (10) losing confidence (11) believe in self-worth (12) general happiness. The same as above, we fit four models with varying degrees of strictness of measurement invariance (see e.g. Kline 2016), namely *configual invariance, weak invariance strong invariance* and *strict invariance* models.

Table A2 below shows the fit measures of these four models. The exact fit hypothesis for the configural invariance is flatly rejected. This implies that the most unconstrained model fits data poorly to start with. This poor fit could be due to very large N, but even other approximate fit measures (CFI, TLI, and SRMR) indicate rather poor fit. This implies that GHQ may not be measuring a one-dimensional concept. A likelihood ratio test rejects the weak invariance model in favour of the configural invariance model (χ2 (22) = 152.86, *P* < 0.001). This again could be due to very large N. In fact, other fit measures of the weak invariance model show an improvement of model fit compared to the configural invariance model. RMSEA, BIC, TLI, and CIT for example favour the weak invariance model. This shows that measurement invariance of GHQ across the three religions can be accepted, however, the unconstrained model fits rather poorly.

**Table A2:**
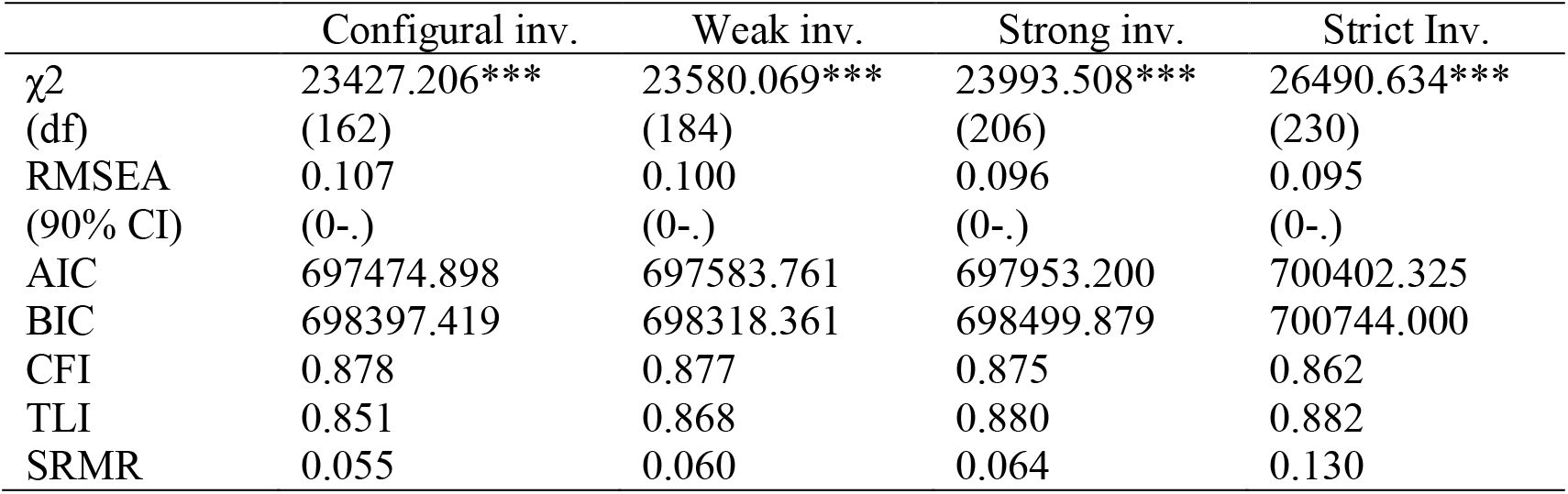
Fit measures of various measurement invariance models of GHQ; N = 37,868.

|  | Configural inv. | Weak inv. | Strong inv. | Strict Inv. |
| --- | --- | --- | --- | --- |
| $\chi^2$ | 23427.206*** | 23580.069*** | 23993.508*** | 26490.634*** |
| (df) | (162) | (184) | (206) | (230) |
| RMSEA | 0.107 | 0.100 | 0.096 | 0.095 |
| (90% CI) | (0-.) | (0-.) | (0-.) | (0-.) |
| AIC | 697474.898 | 697583.761 | 697953.200 | 700402.325 |
| BIC | 698397.419 | 698318.361 | 698499.879 | 700744.000 |
| CFI | 0.878 | 0.877 | 0.875 | 0.862 |
| TLI | 0.851 | 0.868 | 0.880 | 0.882 |
| SRMR | 0.055 | 0.060 | 0.064 | 0.130 |

We now modify the configural invariance model to attain a relatively better fitting baseline model. After inspecting the R-squared per item and the differences in loadings across the three religions, we remove items 1, 3, 4, 7, and 8. The R-squared values for these items are respectively 33%, 23%, 27%, 36%, and 34%. These values are too low to justify including them in the same scale. Next, looking at modification indices, we add an error covariance between item 11 and item 12 and between item 2 and item 5. This results in a relatively well fitting modified configural invariance model with 7 items (see table A3 and Figure A2). We now test various invariance models building on this modified configural invariance model.

**Figure A2:**
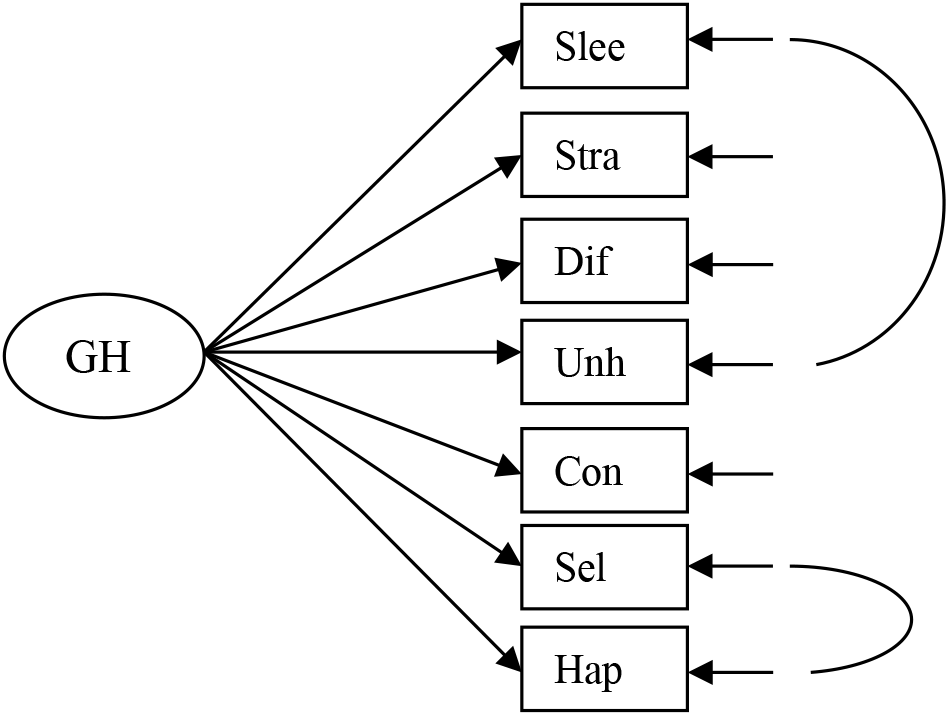
Modified version of GHQ.

The fit of the modified weak invariance model is comparable to the modified configural invariance model. While a likelihood ratio test favours the configural invariance over weak invariance (compare the χ2 values of the two models), this could again be due to very large N. Other fit measures indicate that the modified weak invariance model fits better than the configural invariance model (e.g. RMSEA, BIC, CFI and TLI). Also, in the modified configural invariance model the loadings differ only marginally across the religions. Strong and strict invariance models also fit data relatively well. We thus conclude that the reduced version of the GHQ scale measures wellbeing relatively similarly across the three religions.

**Table A3:** Fit measures of various measurement invariance models of the modified GHQ; N= 37,868.

|  | Configural inv. | Weak inv. | Strong inv. | Strict Inv. |
| --- | --- | --- | --- | --- |
| $\chi^2$ | 2330.605*** | 2395.394*** | 2599.167*** | 3314.358*** |
| (df) | (36) | (48) | (60) | (78) |
| RMSEA | 0.071 | 0.062 | 0.058 | 0.057 |
| (90% CI) | (0.068-0.073) | (0.060-0.064) | (0.056-0.060) | (0.056-0.059) |
| AIC | 471688.735 | 471729.524 | 471909.297 | 472588.488 |
| BIC | 472278.434 | 472216.667 | 472293.884 | 472819.240 |
| CFI | 0.981 | 0.981 | 0.979 | 0.973 |
| TLI | 0.967 | 0.975 | 0.978 | 0.979 |
| SRMR | 0.024 | 0.028 | 0.036 | 0.074 |

#### Effects of religion on modified versions of SWEMWBS and GHQ

Figure A3 shows the effect of religion variables on the modified, reduced versions of GHQ (Fig A2) and of SWEMBWS (items 6 and 7 removed). These effects are similar to the ones with the full versions of GHQ and SWEMBWS. Note that these effects are estimated with a complete-case analysis, hence standard errors tend to be somewhat larger.

**Figure A3:**
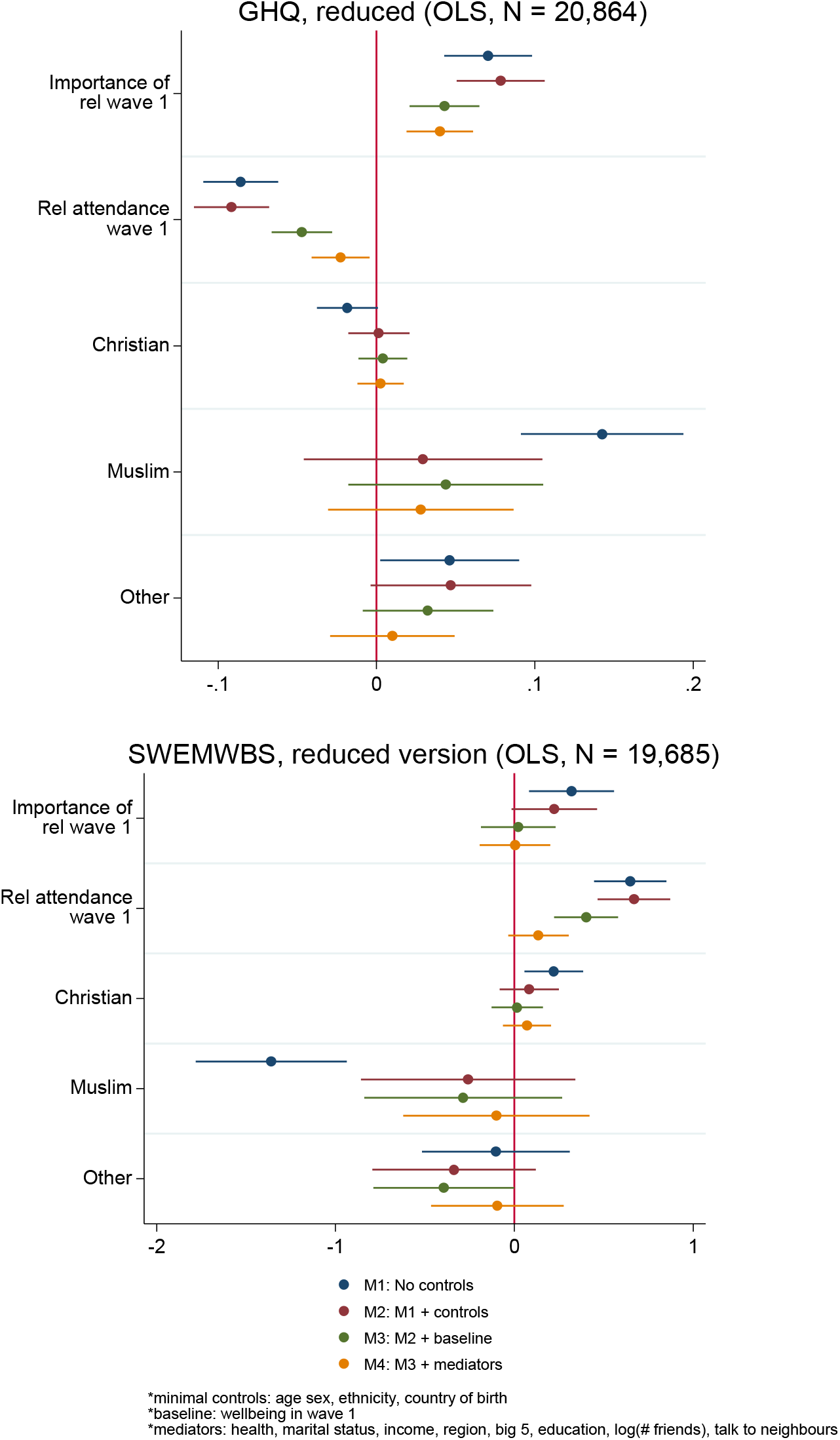
Effects of religion variables on reduced versions of SWEMWBS and GHQ.

### B: Applying survey weights

Here we compare the results with and without survey weights. Note that survey weights are complex when longitudinal and multilevel data are used (see chapter 14 of Snijders and Bosker 2012). We first fit the model without any covariate adjustment with and without the longitudinal adult main survey weights (d_indpxub_lw). Table B1 shows the results. The differences with and without survey weights are minimal, with the exception of the Muslim effect. When covariates are included in the model, the difference between the weighted and unweighted estimates are expected to diminish even further. In fact, we carry out a formal test of interaction between the religion variables and the survey weight using the model with all confounders and mediators as suggested by Snijders and Bosker (2012). The test results shows highly insignificant interaction between the religion variables and the survey weight for both SWEMBWS (χ2 (6) = 6.23, P = 0.398) and GHQ (χ2 (6) = 3.14, P = 0.791). We conclude that applying survey weights or not does not change out conclusions qualitatively.

**Table B1:** Effects of religion variables on SWEMBWS and GHQ with and without survey weights, no covariate adjustment

|  | SWEMBWS |  | GHQ |  |
| --- | --- | --- | --- | --- |
|  | Unweighted | Weighted | Unweighted | Weighted |
| Importance | 0.33**<br>(0.10) | 0.32**<br>(0.12) | 0.66**<br>(0.13) | 0.77**<br>(0.15) |
| Attendance | 0.65**<br>(0.09) | 0.63**<br>(0.10) | -1.03**<br>(0.11) | -1.11**<br>(0.12) |
| Christian | 0.22**<br>(0.07) | 0.24**<br>(0.08) | -0.05<br>(0.09) | -0.01<br>(0.10) |
| Muslim | -1.48**<br>(0.16) | -1.10**<br>(0.21) | 0.96**<br>(0.20) | 0.61*<br>(0.27) |
| Other | -0.24<br>(0.16) | -0.17<br>(0.20) | 0.40*<br>(0.19) | 0.46+<br>(0.24) |
| _cons | 24.20**<br>(0.05) | 24.16**<br>(0.05) | 11.25**<br>(0.06) | 11.22**<br>(0.07) |
| N | 25354 | 22967 | 25252 | 22894 |

Standard errors in parentheses
+ $p < 0.1$ , \* $p < 0.05$ , \*\* $p < 0.01$

### C: Not adjusting for ethnicity

The table below shows the effects of religion variables with and without controlling for ethnicity, which could be colinear with religion.

**Table C1:** Effects of religion variables on SWEMBWS and GHQ with and without adjusting for ethnicity, based on complete case analysis (FIML takes a few hours to converge). All other controls and mediators are included in the models.

|  | SWEMBWS |  | GHQ |  |
| --- | --- | --- | --- | --- |
|  | Adjusted for ethnicity | Unadjusted for ethnicity | Adjusted for ethnicity | Unadjusted for ethnicity |
| Importance | -0.061<br>(0.097) | -0.078<br>(0.096) | 0.463***<br>(0.121) | 0.465***<br>(0.121) |
| Attendance | 0.155+<br>(0.083) | 0.152+<br>(0.083) | -0.307***<br>(0.105) | -0.318***<br>(0.105) |
| Christian | 0.042<br>(0.066) | 0.045<br>(0.066) | 0.024<br>(0.083) | 0.020<br>(0.083) |
| Muslim | -0.254<br>(0.260) | -0.725**<br>(0.192) | 0.204<br>(0.338) | 0.472<br>(0.239) |
| Other | -0.016<br>(0.179) | 0.041<br>(0.168) | 0.184<br>(0.222) | 0.094<br>(0.419) |

Standard errors in parentheses
+ p<0.1, \* p<0.05, \*\* p<0.01

### D: Life satisfaction as the outcome variable

The figure below shows the effects of religion variables on the likert scale measure of life satisfaction measured in wave 4 and wave 2.

**Figure D1.**
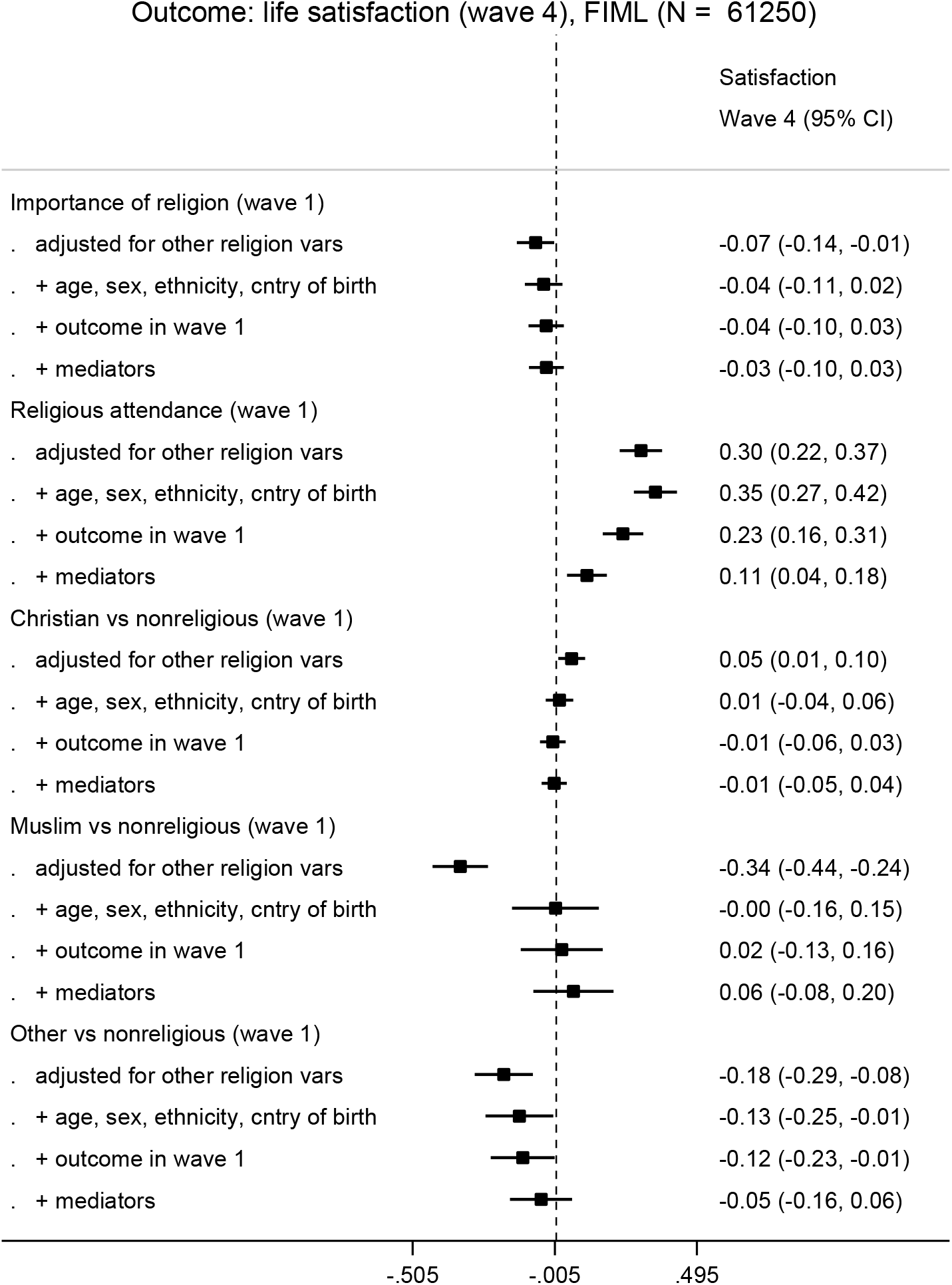

**Figure D2.**
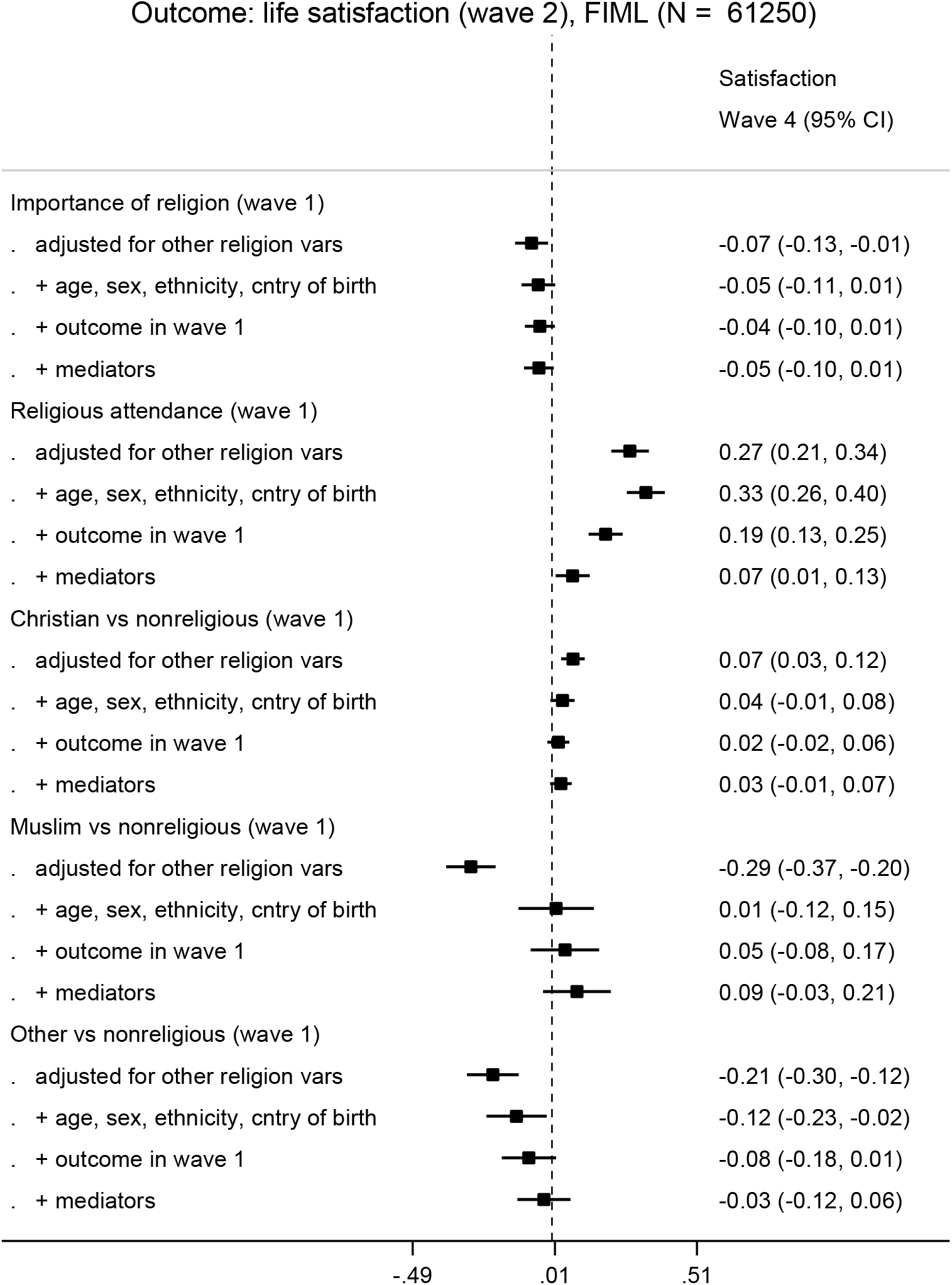

### E: Regressing GHQ measured in wave 2 on religion variables measured in wave 1

The figure below shows the effects when GHQ in wave 2 is regressed on religion variables in wave 2. WEMWBS was not measured in wave 2.

**Figure E1.**
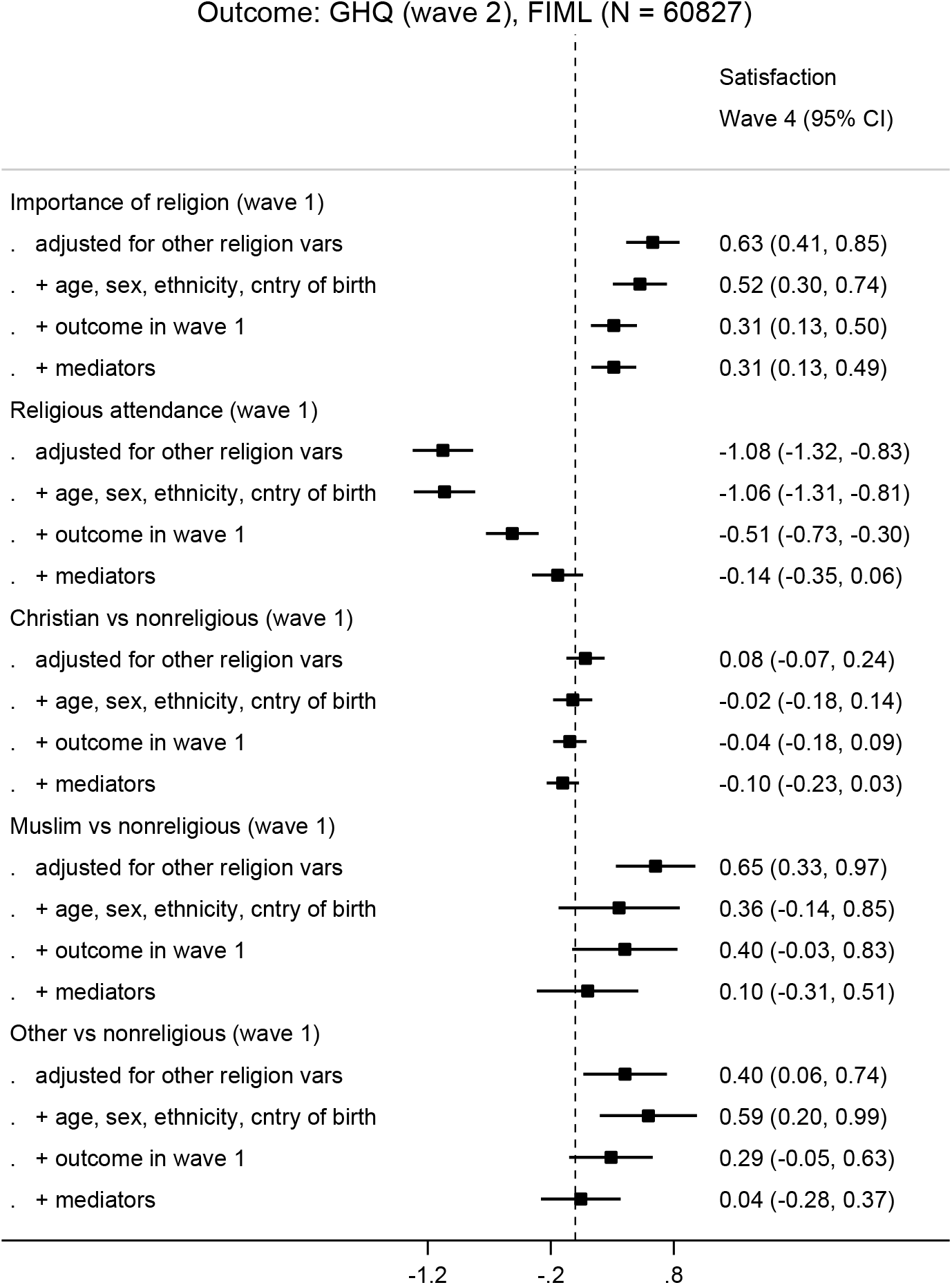

## References

1. Vigo D, Thornicroft G, Atun R. Estimating the true global burden of mental illness. The Lancet Psychiatry 2016;3(2):171–78.

2. Ryan RM, Deci EL. On happiness and human potentials: A review of research on hedonic and eudaimonic well-being. Annu Rev Psychol 2001;52(1):141–66.

3. Steptoe A, Deaton A, Stone AA. Subjective wellbeing, health, and ageing. Lancet 2015;385(9968):640–48.

4. Oswald AJ, Proto E, Sgroi D. Happiness and productivity. Journal of Labor Economics 2015;33(4):789–822.

5. VanderWeele TJ. Religion and health in Europe: cultures, countries, context. Eur J Epidemiol 2017;32(10):857–61.

6. VanderWeele TJ. Religion and health: a synthesis. Spirituality and religion within the culture of medicine: From evidence to practice 2017:357–402.

7. AbdAleati NS, Zaharim NM, Mydin YO. Religiousness and mental health: systematic review study. J Relig Health 2016;10.1007/s10943-014-9896-1

8. Putnam RD, Lim C. Religion, social networks, and subjective well-being. Am Sociol Rev 2010;75(5)

9. Voas D, Chaves M. Is the United States a counterexample to the secularization thesis? American Journal of Sociology 2016;121(5):1517–56.

10. Davey Smith G. Post–Modern Epidemiology: When Methods Meet Matter. Am J Epidemiol 2019;188(8):1410–19.

11. Campante F, Yanagizawa-Drott D. Does religion affect economic growth and happiness? Evidence from Ramadan. The Quarterly Journal of Economics 2015;130(2):615–58.

12. Wallace S, Nazroo J, Bécares L. Cumulative effect of racial discrimination on the mental health of ethnic minorities in the United Kingdom. Am J Public Health 2016;106(7):1294–300.

13. Jordanova V, Crawford MJ, McManus S, et al. Religious discrimination and common mental disorders in England: a nationally representative population-based study. Soc Psychiatry Psychiatr Epidemiol 2015;50(11):1723–29.

14. Louise Casey. The Casey Review: A review into opportunity and integration. In: Department for Communities and Local Government, ed. London, UK, 2016.

15. Meer N, Modood T. Muslim-state relations in Great Britain: an evolving story. Muslim Minority-State Relations: Springer 2016:25–59.

16. Angrist JD, Pischke J-S. Mostly harmless econometrics: An empiricist’s companion: Princeton university press 2008.

17. Kendler KS, Myers J. A developmental twin study of church attendance and alcohol and nicotine consumption: a model for analyzing the changing impact of genes and environment. Am J Psychiatry 2009;166(10):1150–55.

18. Buck N, McFall S. Understanding Society: design overview. Longitudinal and Life Course Studies 2011;3(1):5–17.

19. Stewart-Brown S, Tennant A, Tennant R, et al. Internal construct validity of the Warwick-Edinburgh mental well-being scale (WEMWBS): a Rasch analysis using data from the Scottish health education population survey. Health and quality of life outcomes 2009;7(1):15.

20. Tennant R, Hiller L, Fishwick R, et al. The Warwick-Edinburgh mental well-being scale (WEMWBS): development and UK validation. Health and Quality of life Outcomes 2007;5(1):63.

21. Goldberg DP, Hillier VF. A scaled version of the General Health Questionnaire. Psychol Med 1979;9(1):139–45.

22. Hardy GE, Shapiro DA, Haynes CE, et al. Validation of the General Health Questionnaire-12: Using a sample of employees from England’s health care services. Psychol Assess 1999;11(2):159.

23. Bryan GT, Choi JJ, Karlan D. Randomizing Religion: The Impact of Protestant Evangelism on Economic Outcomes: National Bureau of Economic Research, 2018.

24. Rözer J, Kraaykamp G. Income inequality and subjective well-being: A cross-national study on the conditional effects of individual and national characteristics. Social indicators research 2013;113(3):1009–23.

25. Ngamaba KH, Soni D. Are happiness and life satisfaction different across religious groups? Exploring determinants of happiness and life satisfaction. Journal of religion and health 2018;57(6):2118–39.

26. Helbling M. Opposing muslims and the muslim headscarf in Western Europe. European Sociological Review 2014;30(2):242–57.

27. Ellison CG, DeAngelis RT, Güven M. Does religious involvement mitigate the effects of major discrimination on the mental health of African Americans? Findings from the Nashville Stress and Health Study. Religions 2017;8(9):195.

28. World Health Organization. Handbook on health inequality monitoring: with a special focus on low-and middle-income countries: World Health Organization 2013.

29. Collishaw S. Annual research review: secular trends in child and adolescent mental health. Journal of Child Psychology and Psychiatry 2015;56(3):370–93.

30. Krieger N. Religious Service Attendance and Suicide Rates—Reply. JAMA psychiatry 2017;74(2):197–98.

31. Ross A, Kelly Y, Sacker A. Time trends in mental well-being: the polarisation of young people’s psychological distress. Soc Psychiatry Psychiatr Epidemiol 2017;52(9):1147–58.

32. Naqshbandi M. UK Mosque Statistics/Masjid Statistics, 2017.

